# Cross-Phenotype Plasma Proteomics Reveals Molecular Heterogeneity in Neovascular AMD

**DOI:** 10.64898/2026.05.26.26354036

**Authors:** Rianne Rijken, Els M Pameijer, Aafke de Ligt, Marilette Stehouwer, Saskia M Imhof, Alberta A H J Thiadens, Anneke I den Hollander, Bram Gerritsen, Xuan-Thanh-An Nguyen, Carel B Hoyng, Evianne L de Groot, L. Ingeborgh van den Born, Jeannette Ossewaarde-van Norel, Leonoor I Los, Lude Moekotte, Magda A Smoor, Maria M van Genderen, Ninette H Ten Dam-van Loon, Ramon A C van Huet, Camiel J F Boon, Yvonne de Jong-Hesse, Joke H de Boer, Redmer van Leeuwen, Jonas J W Kuiper

**Affiliations:** Department of Ophthalmology, University Medical Center Utrecht, Utrecht University, Utrecht, the Netherlands; Center for Translational Immunology, University Medical Center Utrecht, Utrecht University, Utrecht, the Netherlands; Department of Ophthalmology, St. Antonius Hospital, Nieuwegein, the Netherlands; Department of Ophthalmology, Erasmus University Medical Center, Rotterdam, the Netherlands; Department of Ophthalmology, Donders Institute for Brain, Cognition and Behaviour, Radboud University Medical Center, Nijmegen, the Netherlands; Ophthalmology Research, Sanofi, Cambridge MA, USA; Department of Ophthalmology, Leiden University Medical Center, Leiden, the Netherlands; The Rotterdam Eye Hospital, Rotterdam, the Netherlands; Department of Ophthalmology, University Medical Center Groningen, University of Groningen, Groningen, the Netherlands; Bartiméus, Diagnostic Centre for Complex Visual Disorders, Zeist, the Netherlands; Department of Ophthalmology, Amsterdam University Medical Center, Amsterdam, the Netherlands

**Keywords:** Targeted proteomics, complement factor H, inherited retinal diseases, circulating inflammatory proteins, multifocal choroiditis

## Abstract

Age-related macular degeneration (AMD) shows substantial clinical heterogeneity that remains unexplained despite extensive genetic and clinical characterization. We evaluated whether proteomic stratification could provide insight beyond clinical phenotype and genetic risk. We performed 384-plex plasma proteomics in a cohort of 215 individuals, including patients with early and late neovascular AMD, other complement-associated retinal diseases, and age-matched controls. Proteome-based reclassification identified four disease-overarching clusters. Neovascular AMD cases were partitioned almost exclusively between two clusters (30/36). Early AMD cases were predominantly assigned to one of these clusters (10/18), whereas only two localized to the other (2/18). Both AMD-associated clusters shared elevated levels of a protein module enriched for lipoprotein-related functions compared to the other clusters. However, the cluster containing both early and neovascular AMD cases showed higher levels of additional protein modules enriched for complement pathways and cellular stress-response pathways compared with the other AMD-associated cluster. Importantly, this molecular divergence in neovascular AMD could not be explained by genetic predisposition (i.e., 52-variant AMD genetic risk score), signatures of biological ageing, nor by other clinical features. Together, these findings support two proteomic endotypes of neovascular AMD with distinct involvement of cellular stress pathways.

## Introduction

Age-related macular degeneration (AMD) is a major cause of visual impairment among adults over 55 years of age^1^. AMD is a clinically heterogeneous disease that varies in presentation, progression rate, and visual outcome. This heterogeneity is evident in its progression through distinct disease stages, from early-to-intermediate (hereafter referred to as early AMD) to late-stage AMD. In early AMD, clinical examination or color fundus imaging shows enlarged lipid-rich deposits under the macula, known as drusen, together with early changes in the retinal pigment epithelium. Many individuals remain stable for years, but the 5-year risk of progression to late AMD ranges from ∼0.3–4% in the lowest risk categories to up to ∼50–70% in the highest risk groups, depending on baseline disease characteristics as well as genetic and lifestyle risk factors^2,3^. Late-stage AMD has two distinct clinical phenotypes: neovascular (“wet”) AMD, which is defined by the presence of neovascularization of the subretinal space, or geographic (“dry”) AMD, which involves degeneration of the photoreceptor and retinal pigment epithelial cells. Intravitreal anti–vascular endothelial growth factor (anti-VEGF) injections remain the most effective treatment to slow or stabilize neovascular AMD, but this treatment does not cure the disease. For geographic (“dry”) AMD, effective treatment options remain limited. Although complement inhibitors targeting C3 and C5 have been approved by the FDA for geography atrophy, they are not approved in Europe due limited efficacy and safety concerns^4–8^. While the AMD phenotype varies across disease stages, considerable heterogeneity also exists within each stage in disease severity, anatomical features, and visual function. Patients with neovascular AMD also vary in their responses to anti-VEGF treatment, indicating potential differences in the underlying disease pathways that drive AMD^9,10^. Understanding these pathways will improve current clinical classification systems and guide the development of new therapies.

Genetic studies have uncovered important insights into AMD genetic predisposition. A genome-wide association study of approximately 16,000 cases and 18,000 controls identified 52 common and rare variants that together explain about half of the genetic predisposition to AMD, forming the basis of the AMD genetic risk score (GRS-52 score)^11,12^. Many of these variants map to genes involved in complement regulation and immune-signaling, with the primary disease association mapping to the Complement Factor H (CFH) gene locus^12–17^. Interestingly, the AMD risk variants in CFH have also recently been implicated in other vision-threatening retinal conditions, such as CRB1-mediated inherited retinal dystrophy (IRD) and multifocal choroiditis (MFC), suggesting that the retina may be particularly sensitive to changes in the complement system^18,19^.

Several studies have linked blood immune protein changes to AMD, including changes in complement factors and cytokines, but they relied mostly on untargeted screens or small panels^20^. Such untargeted approaches are valuable for discovery but often have limited sensitivity and quantification accuracy, making it difficult to capture low-abundance immune proteins consistently. High-throughput targeted proteomics now makes it possible to profile hundreds of circulating proteins with high accuracy and across a large range of concentrations, offering a comprehensive snapshot of the immunological state of an individual. A recent large, targeted proteomics study demonstrated that blood protein profiles can be used to infer the clinical severity of disease and predict progression independent of genetic risk^21^. These findings support the notion that blood-based targeted proteomics may be a promising tool to characterize molecular heterogeneity in AMD and retinal related diseases.

In this study, we applied targeted proteomics to characterize the immunological state of a cohort of AMD and related retinal conditions associated with CFH gene variants. Our objective was to assess whether molecular reclassification based on proteomic profiles can identify distinct clusters of AMD cases with shared pathogenic pathways, and whether this approach provides insights beyond genetic risk scores and clinical phenotypes.

## Results

The study design and main analytic strategies are illustrated in **Figure 1A**. Using state-of-the-art targeted proteomics, we quantified the plasma proteome of 215 high-quality samples across six retinal disease phenotypes: early and late neovascular AMD, *CRB1*-mediated IRD, MFC, and two control groups of different age intervals. After stringent quality control and preprocessing, we obtained a high-quality data set of 368 plasma proteins, including many complement system components, coagulation factors, immune mediators, and lipoproteins. The characteristics of the participants in the retinal phenotype groups are detailed in **Table 1**.

**Table 1.** Clinical and demographic characteristics of the retinal phenotypes. This table summarizes clinical parameters for each of the retinal phenotypes. The distribution of age and disease duration was expressed as median (IQR), while categorical values are reported as counts (n) and percentages (%). Continuous variables were compared across clusters using the Kruskal–Wallis test, and categorical variables using Fisher’s exact test. Abbreviations: N= total; AMD = Age-Related Macular Degeneration; aHC = ageing healthy control; eAMD = early AMD; wAMD = “wet” neovascular AMD; yHC = younger healthy control; IRD= inherited retinal dystrophy; MFC = multifocal choroiditis; F =female; M = Male; CFH = Complement factor H, snv = single nucleotide variant; IQR = interquartile range.

| Characteristic | Overall<br>N = 215<sup>1</sup> | aHC<br>N = 30<sup>1</sup> | eAMD<br>N = 18<sup>1</sup> | wAMD<br>N = 36<sup>1</sup> | yHC<br>N = 26<sup>1</sup> | IRD<br>N = 24<sup>1</sup> | MFC<br>N = 81<sup>1</sup> | p-value<sup>2</sup> |
| --- | --- | --- | --- | --- | --- | --- | --- | --- |
| **Sex** |  |  |  |  |  |  |  | **<0.001** |
| F | 156 (73%) | 20 (67%) | 12 (67%) | 21 (58%) | 13 (50%) | 17 (71%) | 73 (90%) |  |
| M | 59 (27%) | 10 (33%) | 6 (33%) | 15 (42%) | 13 (50%) | 7 (29%) | 8 (9.9%) |  |
| **Age** | 52 (31 – 75) | 74 (71 – 79) | 82 (74 – 84) | 80 (75 – 84) | 28 (25 – 30) | 22 (17 – 33) | 45 (33 – 53) | **<0.001** |
| **CFH snv genotype** |  |  |  |  |  |  |  | **<0.001** |
| 2 | 25 (14%) | 5 (17%) | 0 (0%) | 0 (0%) | 6 (23%) | 13 (54%) | 1 (2.0%) |  |
| 1 | 70 (38%) | 22 (73%) | 6 (33%) | 9 (26%) | 7 (27%) | 5 (21%) | 21 (41%) |  |
| 0 | 88 (48%) | 3 (10%) | 12 (67%) | 25 (74%) | 13 (50%) | 6 (25%) | 29 (57%) |  |
| Unknown | 32 | 0 | 0 | 2 | 0 | 0 | 30 |  |

<sup>1</sup>Categorical: n (%); Continuous: median (IQR).
<sup>2</sup>Categorical: Chi-squared for larger sample sizes/Fisher for smaller sample sizes; Continuous: Kruskal–Wallis.

**Figure 1:**
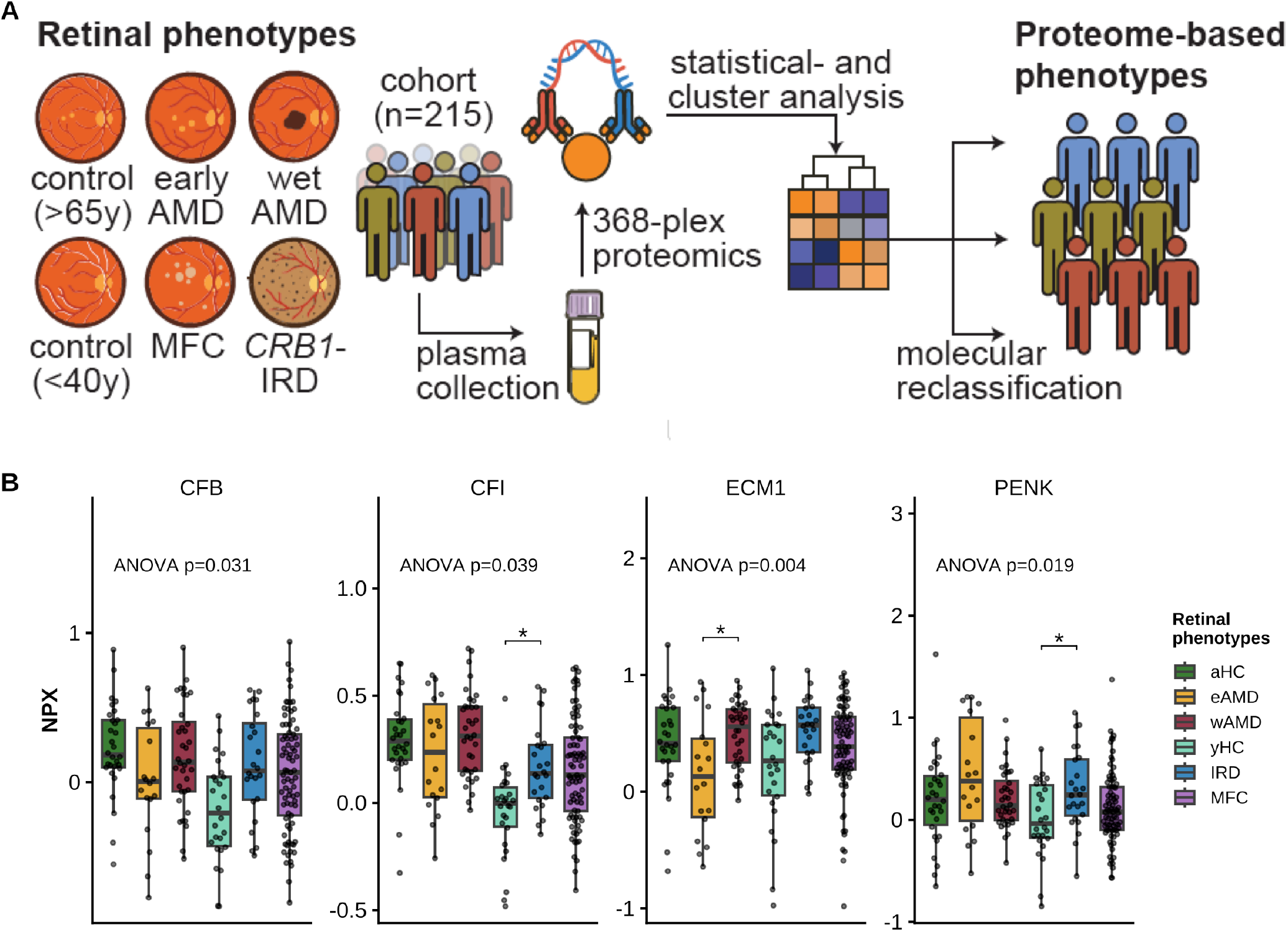
Retinal phenotypes are characterized by diverse plasma proteomes. A) Schematic overview of the study design, showing plasma collection from individuals with retinal phenotypes [early and late wet AMD, multifocal choroiditis (MFC), *CRB1*-associated inherited retinal dystrophy (IRD)], and two control groups [ageing healthy controls (aHC) and younger healthy controls (yHC)], followed by high-throughput proteomic profiling and statistical analysis. B) Boxplots show the four plasma proteins with nominal differential expression between retinal phenotypes. Statistical significance was assessed using ANCOVA adjusted for age and sex, followed by post hoc pairwise comparisons (*nominal P* < 0.05; with * <0.05). None remained significant after correction for multiple testing. Boxplots display normalized protein expression (NPX; log_2_ scale). Horizontal lines indicate the median, boxes represent the interquartile range (IQR), whiskers extend to 1.5× the IQR, and points represent individual samples. Abbreviations: AMD = age-related macular degeneration; eAMD = early AMD, wAMD = late wet AMD; aHC = ageing healthy control; yHC = younger healthy control; IRD = inherited retinal dystrophy; MFC = multifocal choroiditis; NPX = normalized protein expression; ANCOVA = analysis of covariance.

We noted substantial variation in plasma protein levels between patients sharing the same retinal phenotype. Comparison of the levels of individual plasma proteins revealed four proteins with nominal variation between the retinal phenotypes (*nominal P* < 0.05, ANCOVA adjusted for age and sex followed by post hoc pairwise comparisons) (**Supplementary Table 2, Figure 1B**), although none remained significant after correction for multiple testing. Three of these proteins (CFB, CFI, and PENK) differed between the CRB1-mediated IRD patients and the younger healthy controls, whereas extracellular matrix protein 1 (ECM1) showed decreased expression in early-stage AMD compared to both late-stage AMD and the ageing healthy controls.

**Table 2.** Clinical and demographic characteristics of the AMD sample clusters. This table summarizes clinical parameters (age, visual acuity, disease duration) and lifestyle or genetic factors (sex, supplement use, smoking status, early AMD frequency, and genetic risk score [GRS52]) for each of the four proteomic sample clusters (Black, Blue, Green, and Orange) for AMD patient samples. The distribution of age and disease duration was expressed as median (IQR), while categorical values are reported as counts (n) and percentages (%). Continuous variables were compared across clusters using the Kruskal–Wallis test, and categorical variables using Fisher’s exact test. Abbreviations: AMD = Age-Related Macular Degeneration; GRS52 = Genetic Risk Score based on 52 variants; IQR = interquartile range.

| Characteristic | Overall<br>N = 54<sup>1</sup> | Green<br>N = 24<sup>1</sup> | Blue<br>N = 7<sup>1</sup> | Orange<br>N = 18<sup>1</sup> | Black<br>N = 5<sup>1</sup> | p-value<sup>2</sup> |
| --- | --- | --- | --- | --- | --- | --- |
| **Age** | 81 (75 – 84) | 82 (77 – 85) | 77 (63 – 84) | 79 (72 – 83) | 80 (76 – 83) | 0.4 |
| **Worst eye visual acuity** | 0.46 (0.16 – 0.76) | 0.36 (0.20 – 0.74) | 0.58 (0.05 – 1.00) | 0.60 (0.12 – 0.80) | 0.22 (0.16 – 0.32) | 0.5 |
| **GRS52** | 1.86 (1.12 – 2.79) | 1.62 (1.08 – 2.72) | 1.95 (1.24 – 2.57) | 2.39 (1.32 – 2.88) | 1.02 (0.97 – 2.86) | 0.6 |
| **Disease duration (months)** | 70 (22 – 109) | 62 (20 – 87) | 28 (1 – 82) | 81 (34 – 145) | 107 (60 – 387) | 0.11 |
| **Sex** |  |  |  |  |  | 0.3 |
| F | 33 (61%) | 13 (54%) | 4 (57%) | 14 (78%) | 2 (40%) |  |
| M | 21 (39%) | 11 (46%) | 3 (43%) | 4 (22%) | 3 (60%) |  |
| **Supplements use** |  |  |  |  |  | 0.12 |
| No | 17 (31%) | 7 (29%) | 2 (29%) | 4 (22%) | 4 (80%) |  |
| Yes | 37 (69%) | 17 (71%) | 5 (71%) | 14 (78%) | 1 (20%) |  |
| **Early AMD** |  |  |  |  |  | **0.017** |
| No | 36 (67%) | 14 (58%) | 2 (29%) | 16 (89%) | 4 (80%) |  |
| Yes | 18 (33%) | 10 (42%) | 5 (71%) | 2 (11%) | 1 (20%) |  |
| **Smoking status** |  |  |  |  |  | 0.2 |
| Never | 22 (41%) | 8 (33%) | 5 (71%) | 8 (44%) | 1 (20%) |  |
| Former | 27 (50%) | 11 (46%) | 2 (29%) | 10 (56%) | 4 (80%) |  |
| Current | 5 (9.3%) | 5 (21%) | 0 (0%) | 0 (0%) | 0 (0%) |  |

<sup>1</sup>Categorical: n (%); Continuous: median (IQR).
<sup>2</sup>Categorical: Chi-squared/Fisher as appropriate; Continuous: Kruskal–Wallis.

### Molecular reclassification identifies disease-overarching sample clusters across retinal phenotypes

Heterogeneity in plasma proteins between patients within the same retinal phenotype might suggest that different disease pathways lead to the same retinal phenotype. If this is indeed the case, molecular reclassification of samples based on their plasma proteomes could identify patient clusters that share molecular mechanisms (i.e., molecular endotypes). We aimed to accomplish this by resampling-based consensus clustering to identify stable clusters of samples and robust co-expressed protein modules. This resulted in four sample clusters of comparable size (n = 44–66), and four modules of co-expressed plasma proteins (**Figure 2**). We noted a significant difference in age between the sample clusters (**Supplementary Table S3**), with the *Green* and *Orange* clusters showing comparable mean age, as well as the *Black* and *Blue* clusters. As expected, comparison of individual plasma protein levels adjusted for age and sex revealed substantial variation between the sample clusters (n = 310, *Padj* < 0.05, ANCOVA adjusted for age and sex followed by post hoc pairwise comparisons) (**Supplementary Table S4**).

**Figure 2:**
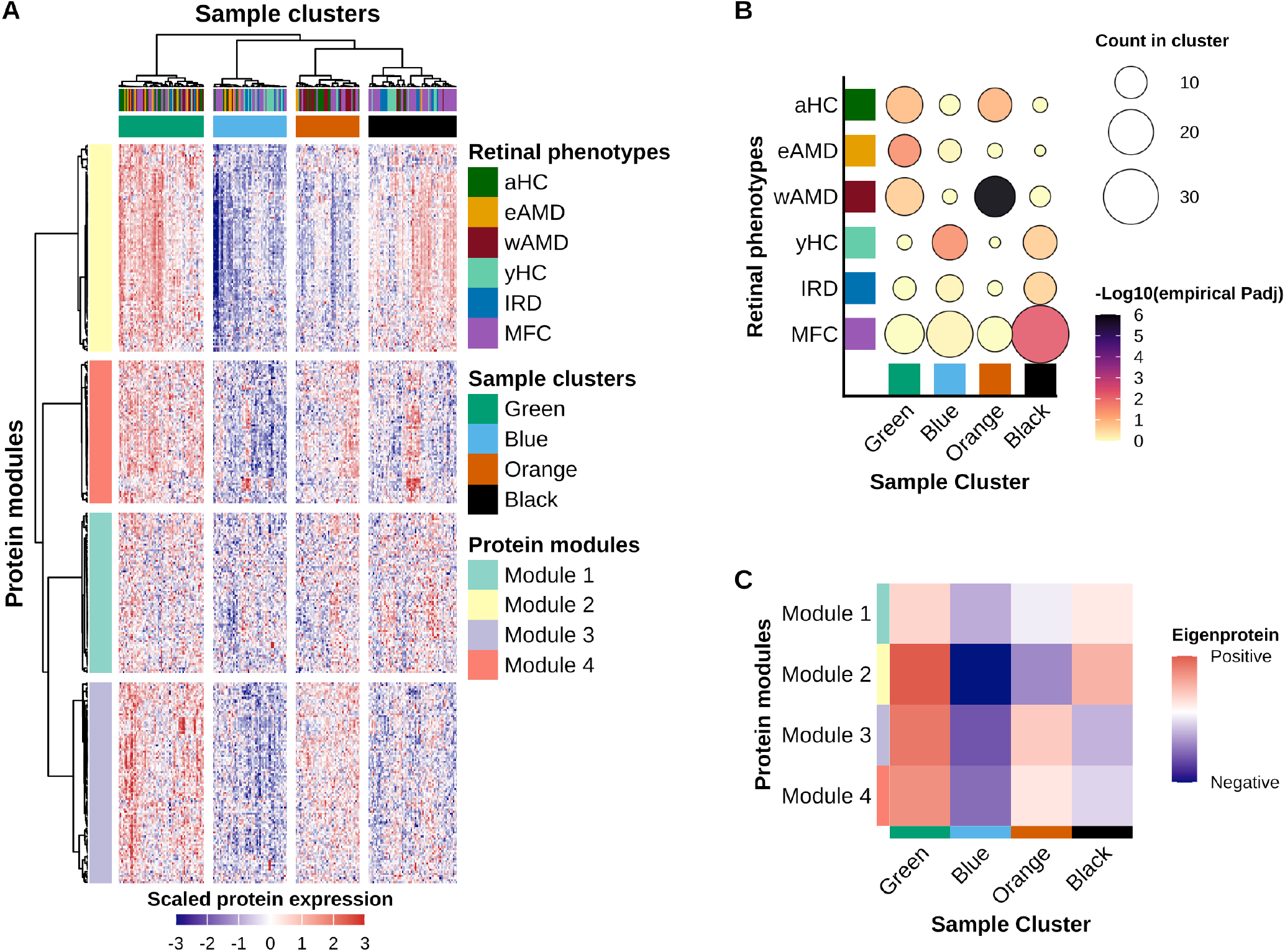
Molecular reclassification of the plasma proteomes identifies four disease-overarching signatures. A) Heatmap shows the consensus clustering of the cohort. Columns represent the four sample clusters (*Black, Orange, Blue, Green*), annotated with their corresponding retinal phenotypes. Rows represent the four protein modules (Module 1-4) and clustering results are shown by the dendrograms. B) Overrepresentation analysis shows the relative enrichment of clinical retinal phenotypes within each sample cluster. Enrichment was tested using 2,000-permutation empirical p-values, corrected for multiple testing (FDR). Bubble size indicates the number of samples per phenotype, and color intensity represents the statistical significance (–log_10_(FDR)). C) Heatmap is showing the Eigenprotein values of the protein modules expressed across the sample clusters. The module Eigenprotein represents the first principal component summarizing the coordinated expression of proteins within each module. Positive Eigenprotein values (red) indicate higher average expression of the module proteins in that cluster, while negative Eigenprotein values (blue) indicate lower average expression. Abbreviations: aHC = ageing healthy control; eAMD = early age-related macular degeneration; wAMD = late wet age-related macular degeneration; yHC = younger healthy control; IRD = inherited retinal dystrophy; MFC = multifocal choroiditis; Padj = adjusted p-value; FDR = false discovery rate.

### Genetic variation explains only a minor portion of the sample clustering

Because a common polymorphism within the *CFH* gene is linked to AMD, MFC and IRD, we tested whether the proteomic clusters could be attributed to the CFH genotype^12,18,21–24^. After adjusting for *CFH* genotype using a likelihood ratio test, 23 proteins showed nominal significant differences, but only CFHR2 remained significant after correction for multiple testing (*Padj* = 1.25 × 10-5). This finding indicates that only a minor portion of the differences between the sample clusters can be attributed to the *CFH* genotype (**Supplementary Table S5**). AMD genetic risk scores (i.e., GRS-52 scores) were only available to AMD patients, but this genetic score also did not significantly differ between AMD patients assigned to different clusters. This indicates that AMD risk variants do not account for the bulk of the proteomic differences between the sample clusters (**Table 2**).

### Proteomic sample clusters differ in their enrichment of clinical retinal phenotypes

We subsequently tested whether each sample cluster showed concordance with the clinical retinal phenotypes. Sample distribution was relatively balanced across clusters (44–61 samples each; **Supplementary Table S3**). Although all clusters contained more than one phenotype, the permutation-based overrepresentation analysis showed significant enrichment after FDR-correction (**Figure 2B**; **Supplementary Table 6**).

The *Green* and *Orange* clusters represented older individuals (median age 73 years) and were dominated by AMD and ageing control cases and some of the MFC cases (**Supplementary Table S3**). The *Green* cluster, which contained the largest proportion of AMD cases (24 of 59 samples) including part of the neovascular cases (n=14) and about half of early AMD cases (n=10), showed a significant enrichment for early AMD (empirical *Padj* = 0.036) (**Table 2**; **Supplementary Table 6**). The *Orange* cluster, which also included AMD cases (18 of 44 samples) together with the older healthy controls, was strongly enriched for neovascular AMD (16 cases; empirical *Padj* < 0.001). The younger clusters, *Black* (median age 29 years) and *Blue* (median age 38 years), included many MFC samples, and most of IRD cases and nearly all younger control samples. The *Black* cluster showed a significant enrichment for MFC (33 cases; empirical *Padj* = 0.006), while the *Blue* cluster was significantly enriched for younger controls (12 cases; empirical *Padj* = 0.036). The *Blue* cluster also contained five early AMD cases, representing about one-quarter of all early AMD samples. IRD did not show significant enrichment in any sample cluster. Together, the reclassification in sample clusters shows that patients within the same retinal phenotype can differ in their proteomic profiles, meaning that proteomic profiles capture heterogeneity that is not visible from clinical observations alone.

### Protein modules reveal shared biological signatures within sample clusters

Next, we investigated the profile of protein modules that defined the sample clusters. Pathway enrichment analysis indicated that Module 1 (n = 78 proteins) was enriched for Fibroblast Growth Factor Receptor 4 (FGFR4)-signalling, Module 2 (n = 100) for stress response pathways, Module 3 (n = 97) for lipoprotein-related functions, and Module 4 (n = 69) for the complement cascade (**Supplementary Figure S2B**). We then summarized each protein module using the module Eigenprotein (first principal component of each protein module) and linked these to the sample clusters (**Figure 2C**; **Supplementary Figure S2A**). The *Green* cluster contained the largest number of AMD cases and exhibited elevated levels of modules 2, 3, and 4. The *Orange* cluster maintained positive expression of modules 3 (lipoprotein) and 4 (complement), but these levels were clearly lower than in the *Green* cluster, whereas module 2 (stress pathways) was substantially reduced. The *Blue* cluster, composed largely of younger controls, displayed lower eigenprotein values across all modules, indicating comparatively low levels of acute-phase proteins, complement components, and other immune factors. In contrast, the *Black* cluster, dominated by MFC cases, showed increased levels of module 2, but did not exhibit the elevation in the lipoprotein module 3 or complement module 4 proteins linked to clusters with the most AMD cases. Taken together, these protein modules patterns show that the *Green* cluster represents the most immune-active AMD subgroup, while the *Orange* cluster reflects a relatively more moderate immune phenotype. The *Blue* and *Black* clusters have a distinct proteomic profile, suggesting differences in underlying molecular signaling pathways for MFC and controls.

### Correlation-based network analysis identifies hub proteins representing core biological processes

To better understand the underlying biology of the differences in protein modules between the samples clusters, we then constructed a correlation-based protein network to obtain hub proteins (i.e., proteins most strongly correlated with the module Eigenprotein, in kME) that are likely to drive the biological processes associated with the plasma protein modules (**Figure 3A**;**Supplementary Table 7**). To explore how these hub proteins varied across retinal phenotypes, their z-scored expression levels were examined across sample clusters (**Figure 3B**), revealing coordinated, module-specific expression patterns. Representative hub proteins for each module differed significantly between sample clusters (**Figure 3C**), indicating that these hub proteins reliably capture module-level behavior. The lipoprotein-related module 3, which was associated with both AMD clusters (Green and Orange), included ADAMTS1 (a disintegrin and metalloproteinase with thrombospondin type 1 motif 1) and complement factor D (CFD, also known as adipsin) among its hub proteins. The complement-enriched module 4, linked to both the *Green* AMD cluster and the MFC-dominated *Black* cluster, contained the hub proteins C1S (a component of the classical complement pathway) and complement factor I (CFI), a complement convertase inhibitor. In contrast, module 2, which distinguished neovascular AMD cases (in *Green* and *Orange* cluster), was centered on the hub proteins MARS1 (methionyl-tRNA synthetase 1) and VTI1A (vesicle transport through interaction with t-SNAREs 1A).

**Figure 3:**
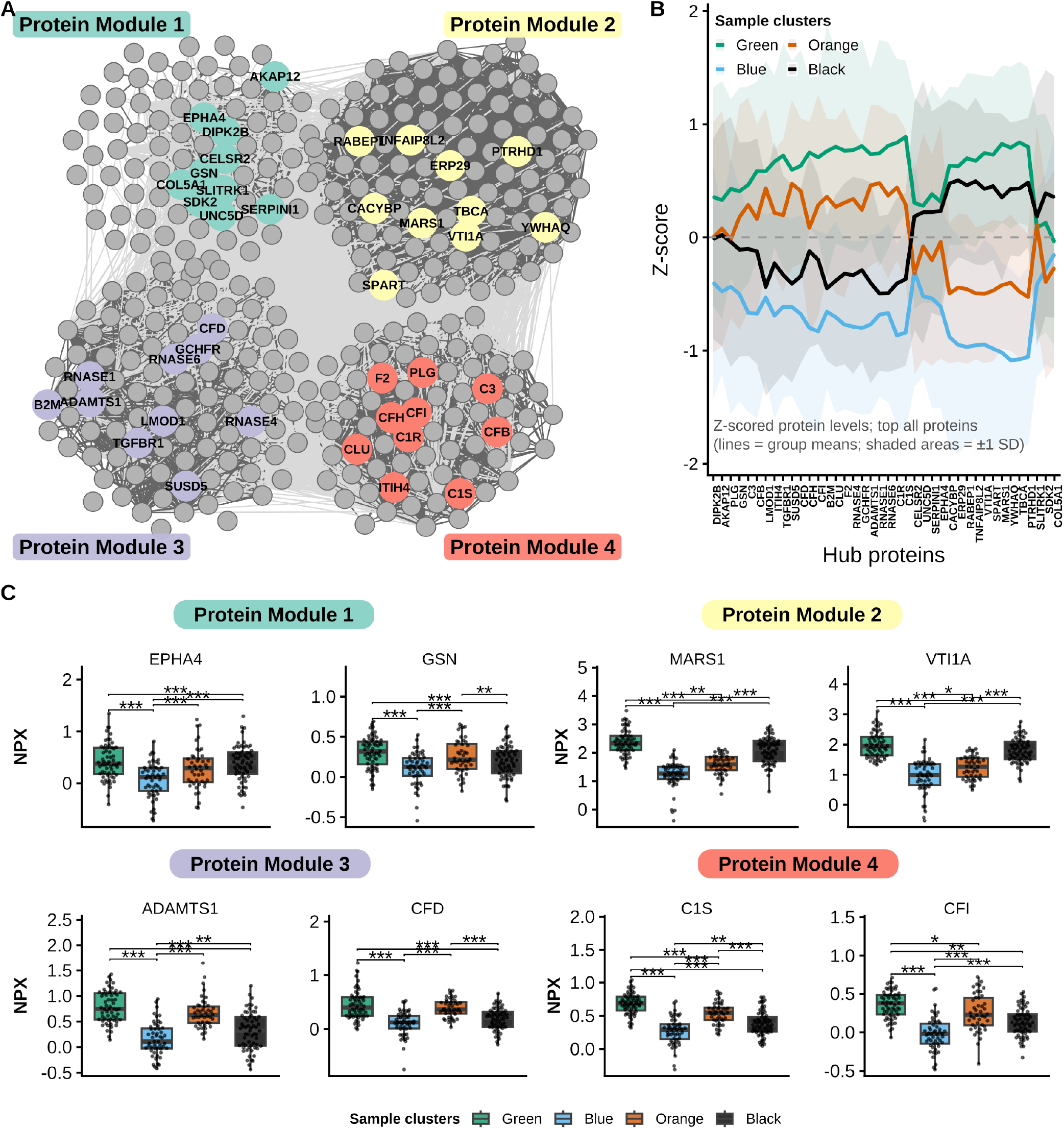
Network-based identification of co-expressed plasma protein modules and their differential expression across sample clusters. A) Correlation network shows co-expressed protein modules. Top 10 proteins with highest module membership (kME, or Protein centrality within a module) are highlighted, while other proteins are shown in grey. Edges represent significant correlations within modules (darker) or between modules (lighter). B) Parallel-coordinates fingerprint shows 40 DEPs (top 10 proteins with kME per module) across retinal phenotypes ordered by the similarity of mean profiles. Lines (and ribbons) show z-scored group mean(±1 SD), where z-score=0 represents the average of all groups. For each protein we computed the mean z-score per group, correlated proteins (Spearman’s ρ) across groups and calculated distance (1-ρ)/2 followed by hierarchical clustering. C) Boxplots show representative hub proteins from each module: EPHA4, GSN (Module 1); MARS1, VTI1A (Module 2); CFD, ADAMTS1 (Module 3); and C1S, CFI (Module 4). Statistical significance was determined using ANCOVA adjusted for age and sex, followed by post hoc pairwise testing with correction for multiple testing (*Padj* < 0.05; with * < 0.05, ** < 0.01, *** < 0.001). Boxplots display normalized protein expression (NPX; log_2_ scale). Horizontal lines indicate the median, boxes represent the interquartile range (IQR), whiskers extend to 1.5× the IQR, and points represent individual samples.Abbreviations: AMD = Age-related macular degeneration; kME = module membership; DEPs = differentially expressed proteins; NPX = Normalized Protein Expression (log_2_ scale); SD = standard deviation; IQR = interquartile range; ANCOVA = analysis of covariance;Padj = adjusted p-value; ρ = Spearman’s rank correlation coefficient; EPHA4 = Ephrin type-A receptor 4; GSN = Gelsolin; MARS1 = Methionyl-tRNA synthetase 1; VTI1A = Vesicle transport through interaction with t-SNAREs 1A; CFD = Complement factor D; ADAMTS1 = A disintegrin and metalloproteinase with thrombospondin motifs 1; C1S = Complement C1s; CFI = Complement factor I.

### Clinical differences do not explain the proteomic differences between the sample clusters

Finally, we were interested to determine if there were clinical differences that could explain the proteomic differences between the neovascular AMD cases in the *Green* and *Orange* sample clusters. Using available data, we found that the cases in the *Green* and *Orange* sample clusters did not differ in age, visual acuity, disease duration, or genetic predisposition to AMD (i.e., GRS-52 score) (*P* > 0.05, **Table 2**). The proportion of participants using any AMD-related nutritional supplement (AREDS, AREDS-2, Nutrof Omega, or other formulations) did not differ significantly between clusters. Finally, we investigated whether the differences between the two neovascular AMD clusters were driven by proteomic signatures associated with advanced biological ageing, a known risk factor for AMD^25^. As described above, both clusters showed elevated levels of the lipoprotein-related protein module 3. This module was strongly enriched for proteomic signatures linked to peak biological ageing at age 67 (72 of 97 proteins overlapping, one-sided hypergeometric test, *Padj* = 8.73 × 10-6) (**Supplementary Figure S3**). In contrast, the separation between the neovascular AMD clusters was primarily driven by protein module 2 (and potentially module 4), but neither module 2 nor module 4 showed significant enrichment for ageing-associated proteomic signatures (*Padj* > 0.1), indicating that the observed cluster differences cannot be attributed to differences in biological ageing alone.

## Discussion

We investigated the plasma proteome in AMD and other complement-related retinal phenotypes, and identified disease-overarching sample clusters and heterogeneity in neovascular AMD that could not be attributed to genetic background, or other factors implicated in AMD.

Plasma proteomic profiles were largely similar between retinal phenotype groups and controls with only a few nominal differences that did not remain significant after correction for multiple testing. We note that substantial within-group variation limited the power to detect between-group differences. Regardless, extracellular matrix protein 1 (ECM1), a structural component of the extracellular matrix, was significantly lower in early AMD compared to late AMD and age-matched controls, which suggest early changes in remodeling of the extracellular matrix. In AMD, the balance between extracellular matrix production and degradation is disturbed, which may weaken Bruch’s membrane structure in the early disease stage^26,27^. Reclassification of samples based on their proteomic profiles revealed that patients with clinically similar retinal phenotypes can differ substantially in their proteomic profiles. This analysis also subdivided the bulk of neovascular AMD cases into two distinct clusters: one with elevated levels for proteins related to cellular stress pathways, lipoprotein-related processes, and proteins of the complement cascade, while the other cluster showed moderate elevation of lipoprotein- and complement-related protein modules, without concurrent upregulation of cellular stress pathway proteins (module 2). About half of the early AMD cases were found in the cluster with cellular stress pathway proteins. It will be interesting to determine in future studies if they show altered disease course (i.e., more prone to progress to late stage disease) and/or differential therapeutic response. The presence of both early and neovascular AMD cases in this cluster suggests that this molecular profile may span multiple disease stages, although its relationship with disease progression remains unclear.

Elevated levels of complement proteins in AMD, including C3, CFB, CFH, CFI, and C5 (hub proteins of module 4) support complement dysregulation in AMD, which is consistent with known genetic risk associations for AMD^12^. Several of the complement components in this module have also been explored as therapeutic targets. For example, C1S has been linked with retinal inflammation in AMD, but available anti-C1s antibodies have not yet been tested in AMD to date^28^. Reduced CFI activity is linked with increased complement activation and higher AMD risk, but CFI-targeting therapies so far have failed to slow progression of geographic atrophy in AMD^29–31^. Other complement-directed therapies so far struggle to prove discernable clinical impact, which has contributed to regional differences in market access driven by regulatory concerns regarding efficacy and safety^32,33^. Our findings indicate that the limited efficacy observed in these clinical trials could potentially in part be due to pronounced variability in complement component levels between AMD patients. This variation could lead to masked or reduced treatment responses in trials and signal the need of individualized dosing strategies. It would be interesting to determine whether stratifying patients by the plasma levels of key proteins in this module (i.e., CFI or C1S profiles) in historic trial data or future trials may lead to different outcomes in complement-targeted therapies.

The disease-overarching design of our study demonstrated that the cellular stress-related protein module (module 2) also characterizes another cluster of retinal phenotypes that were largely non-AMD (black cluster). Notably, samples in this cluster did not show the AMD-associated complement and lipoprotein protein signatures (module 3 and 4). Because this cluster was enriched for MFC, a highly inflammatory eye disease that shares choroidal neovascularization with neovascular AMD, these findings raise the possibility that this module captures pathogenic mechanisms common to both conditions. The stress-related pathways of this module include oxidative stress mechanisms, which form a well-known risk factor for AMD and are a key driver of angiogenesis, as well as biological aging^34,35^. Therapeutic strategies targeting oxidative or cellular stress pathways may be particulary relevant for this subgroup of cases, such as antioxidant supplementation by Age-Related Eye Disease Study (AREDS) supplements.

Because hierarchical clustering is inherently unsupervised, it does not account for relevant covariates linked to AMD, such as age. Consequently, differences in chronological age between clusters may contribute to the observed proteomic variation. Indeed, large-scale targeted plasma proteomic studies have consistently linked biological ageing and age-related diseases to inflammatory pathways^36^. Our results are consistent with this framework, because the chronologically older *Green* and *Orange* clusters (containing the majority of AMD cases and age-related controls) were characterised by elevated levels of the lipoprotein-related protein module 3, which was strongly enriched for proteomic signatures associated with accelerated biological ageing^25^. In contrast, the cellular stress pathways-linked protein module 2 was not enriched for ageing-associated signatures but was more influential in distinguishing between the *Green* and *Orange* clusters. This indicates that differences in cellular stress pathways between these neovascular AMD subtypes may reflect the contribution of other (environmental or lifestyle) factors that likely act synergistically with ageing-related processes (module 3) and underlying genetic predisposition (GRS-52). Also, few AMD patients were assigned to the *Blue* cluster, which exhibited the lowest levels of the proteins profiled in this study. This pattern may represent either a genuine low-expression disease signature or an alternative disease course driven by proteins not included in the current assay. Additional proteomic profiling may therefore be required to uncover potential proteins characteristic specific to this subgroup.

Several limitations of this study should be considered. A common challenge in disease-overarching studies is the inclusion of samples collected across different sites and protocols and assays across multiple plates, despite randomization of samples across plates. While we accounted for batch and plate effects, this may also have attenuated potential biological differences between retinal phenotypes. In future studies shared reference samples (bridging controls) would be an exciting opportunity to better account for these technical limitations^25^. However, since consensus clustering is based on correlation patterns of scaled data rather than absolute protein levels, the resulting sample clusters and protein modules are more robust against batch correction and technical variation. A second limitation of this study is the cross-sectional design. Longitudinal sampling will be essential to monitor the circulating immune-related proteins in AMD patients over time. This is particulary relevant for tracking cases with early AMD to late AMD. Ideally, future studies would also monitor the blood of healthy people with a family history of AMD and a known genetic risk. Since we identified some plasma immune biomarkers, tracking their levels on several time points could help understand whether inflammation predicts disease progression or therapy responses. Expanding these retinal disease cohorts will help confirm the protein signatures we observed and clarify their relevance across various patient groups and disease stages.

In conclusion, plasma proteomics revealed heterogeneity in the plasma proteins of AMD patients with distinct complement-, and lipid metabolism and cellular stress-pathways involvement that were not explained by clinical retinal phenotype, genetics, or biological ageing. The cluster-specific activity of these pathways may contribute to differences in disease mechanisms and treatment outcomes and supports the potential of circulating protein profiles for patient stratification in retinal diseases.

## Methods

### Study design

The study population comprised a retinal phenotype cohort consisting of clinically distinct phenotypes, including AMD, IRD, and MFC, and controls.

### Ethics

The study was approved by the local Institutional Review Board of the University Medical Center Utrecht (UMCU) (protocols #14–065, #20–428) see also^18,19^.

#### Retinal phenotype cohort

We recruited 60 AMD patients at the outpatient clinics of St. Antonius Hospital and the University Medical Center Utrecht, The Netherlands. All AMD cases and controls were classified using fundus photography and optical coherence tomography, in accordance with the simplified AREDS criteria. Patients with small to intermediate drusen, retinal pigment epithelium (RPE) abnormalities, and pigmentary changes were classified as early AMD (n=21), while all late AMD cases (n=39) had neovascular AMD, as no geographic atrophy cases presented during recruitment. In addition, we recruited 39 age- and sex-matched controls without AMD or other inflammatory retinal phenotypes. The controls were patients referred to the St. Antonius Hospital or UMCU with unrelated ophthalmic conditions. These included eyelid disorders (n = 6), corneal and lens disorders such as Fuchs’ corneal dystrophy and senile cataract (n = 20), and non-inflammatory retinal conditions, including glaucoma suspects, macular hole, vitreomacular traction, and vitreous hemorrhage (n = 13). These were grouped together as ageing controls (aHC). None of the AMD patients and controls had used systemic immunomodulating therapy, had a history of systemic inflammatory diseases, cancer, diabetes, diabetic retinopathy, retinal detachment, or other inflammatory retinal diseases. Samples from 30 CRB1-associated IRD patients were collected, who were characterized by ophthalmic examination, imaging, full-field electroretinography, and genetic confirmation of two or more rare functional variants affecting both gene copies of the CRB1 gene, as we described previously^19^. Patients with systemic inflammatory conditions or systemic immunomodulatory treatment at the time of sampling were excluded. In addition, age-matched younger healthy controls (yHC) were obtained from anonymous blood bank donors (n=29) without a history of inflammatory disease at the University Medical Center Utrecht Research Blood Bank. Samples from 87 MFC patients were collected from individuals recruited and diagnosed according to the Standardization of Uveitis Nomenclature (SUN) Working Group guidelines after a standard diagnostic workup for uveitis, as we described previously^18^. Patients with systemic inflammatory conditions or systemic immunomodulatory treatment at the time of sampling were excluded.

### Blood sample processing

Venous blood was collected into ethylenediaminetetraacetic acid (EDTA) vacutainers (#362084, BD Biosciences, Franklin Lakes, USA) (Supplementary Table S1, *key resources table*). Blood samples were centrifuged at 400 × g for 10 minutes to separate plasma from blood cells. The resulting plasma was transferred to 15 mL tubes and centrifuged again at 1500 × g for 10 minutes. The cleared plasma was aliquoted into Micronic 1.4 mL round-bottom tubes (#MP32022, Micronic, Lelystad, the Netherlands) and stored at –80°C until proteomic analysis. After plasma removal, peripheral blood mononuclear cells (PBMCs) were isolated from the remaining blood fraction using Ficoll density gradient centrifugation, followed by DNA extraction for genotyping as previously described^37^.

### Targeted plasma proteomics profiling

Plasma samples from all cohorts were subjected to targeted proteomic analysis. The samples were randomly distributed across three 96-well plates (#72.1980; Sarstedt, Nümbrecht, Germany) to minimize potential plate-related batch effects. The plates were sealed with adhesive film (#232698; Thermo Fisher, Waltham, MA, USA) and shipped on dry ice to the Olink Proteomics facility at Erasmus Medical Center in Rotterdam, the Netherlands. In total, 368 inflammation-associated plasma proteins were quantified using the Olink® Explore 384 Inflammation II panel. Protein quantification was based on Olink’s proximity extension assay technology combined with next-generation sequencing. Data was reported as normalized protein expression (NPX) values on a log2 scale, where a 1 NPX difference corresponds to a doubling of protein concentration.

### Genotyping AMD risk variants

The AMD cohort was previously genotyped^37^ for 52 common and rare AMD-associated genetic variants using a custom SNP array, as described in a previously established method^11^. The resulting 52 variant genetic risk score (GRS)-52^37^ ranged from – 1.93 to 3.59, with values classified as low (<–0.057), medium (≥–0.057 and <1.131), or high (≥1.131)^38^. The genotype of the CFH variant rs10922109 was obtained from the array data for AMD cases and ageing controls. For the IRD and MFC cohorts, genotype of this variant (proxy rs7535263; r^2^ = 0.98 in EUR superpopulation of the 1000 Genomes) was obtained by genotyping with Infinium OmniExpress-24 bead chip (Illumina) or Taqman genotyping assays, as we described previously^18,19^. Genotypes for rs10922109 (proxy rs7535263) were converted to dosage of the A allele (G/G = 0 (high risk), G/A = 1 (heterozygous), and A/A = 2). Imputed genotype data were rounded to the nearest integer (0, 1, or 2).

### Data processing and quality control

All analyses were performed in R version 4.4.3 (2025-02-28)^39^ with RStudio (2026.4.0.526)^40^. The R packages *rmarkdown*^41^, *tidyverse*^42^, *dplyr*^43^, and *stringr*^44^ R packages were used for data analysis and *ggplot2*^45^, *ggpubr*^46^, *ggrepel*^47^, and *cowplot*^48^ for visualization of the data. Data import and tabular outputs were handled using the R packages readxl^49^, gtsummary^50^, and flextable^51^. Manuscript generation and citation management were performed using officedown^52^, knitr^53^, and citr^54^. Raw NPX data was inspected using the *olink_qc_plot()* and *olink_dist_plot()* from the *OlinkAnalyze*^55^ package, and Principal Components Analysis (PCA) visualization using *prcomp()* from the *stats*^39^ package and density plots. Measurements for the protein TNFSF9 contained 13 missing values and was therefore removed. Proteins with more than 80% of values below the limit of detection (LOD; n = 22) and proteins with assay-level warnings reported by Olink (n = 10) were flagged but retained for downstream analyses and interpreted with caution. At the sample level, one outlier was removed based on its NPX median versus interquartile range (**Supplementary Figure S1A**). In addition, samples with QC-level warnings reported by Olink (n = 28), defined as NPX values deviating more than ±0.3 from the plate median, were excluded (**Supplementary Figure S1B**), leaving 215 high-quality plasma samples for analysis. PCA of these 215 samples revealed a freezing-protocol effect and a plate-batch effect (**Supplementary Figure S1**C–D) that was corrected with batch correction using *ComBat()* from the sva^56^ package. After batch correction, PCA showed that the batch effects had been effectively removed (**Supplementary Figure S1E–F**). These adjustments resulted in a batch-corrected dataset of 215 samples and 368 proteins, which was then used for downstream differential expression analysis. Gene symbols are used to name proteins (e.g., CFH for factor H) and are written in regular (non-italic) font when referring to the protein and in italics when referring to the corresponding gene.

### Statistical analysis

Differential expression analysis was performed using *olink_anova()* and *olink_anova_posthoc()* from the *OlinkAnalyze*^55^ package, adjusting for age and sex. *P*-values were corrected for multiple testing using the Benjamini–Hochberg (BH) false discovery rate (FDR) method^39^. Unsupervised clustering was performed using the *ConsensusClusterPlus*^57^ package. Before clustering, protein expression levels were z-scored per protein across samples. Consensus clustering was applied to both samples and proteins using Euclidean distance and Ward.D2 linkage. The optimal number of sample clusters (k = 4) and protein modules (k = 4) was determined based on evaluation of the consensus cumulative distribution function and delta area plots generated by *ConsensusClusterPlus*^57^.

The influence of the *CFH* genotype on the sample clusters was evaluated using a likelihood ratio test of age- and sex-adjusted models conducting differential expression analysis between the sample clusters with and without *CFH* genotype as a covariate.

Overrepresentation analysis of retinal phenotypes within the sample clusters was performed using a hypergeometric test (equivalent to a one-sided Fisher’s exact test for enrichment). Empirical *P* values were calculated using 2,000 phenotype-label permutations, followed by FDR correction.

The module Eigenprotein was calculated using the first principal component of the standardized expression matrix of all proteins within a module. The kME (module membership) for each protein was then computed as the Pearson correlation between its expression profile and the corresponding module Eigenprotein. Finally, correlation-based network analysis was performed within protein clusters to explore co-regulation patterns and identify hub proteins (proteins with the highest kME values). We visualized the discriminative proteomic signature across retinal phenotypes using a parallel-coordinates approach. For each protein, we calculated the group-wise mean z-score. To order proteins by similarity of expression patterns, we correlated group means between proteins using Spearman’s ρ, transformed the correlations to distances (1 – ρ)/2, and applied hierarchical clustering with average linkage. Functional pathway enrichment was performed using the *clusterProfiler* and *ReactomePA*^58,59^ packages, based on Gene Ontology (GO), Kyoto Encyclopedia of Genes and Genomes (KEGG), and Reactome databases, and visualized using dot plots and heatmaps of −log_10_-transformed adjusted *P*-values. Enrichment analysis for biological ageing-signature as described by Ma et al. 2026 was conducted using a one-sided hypergeometric overrepresentation test, using the full set of module proteins from our study as the background^25^.

## Data availability

The datasets generated and/or analysed during the current study are available in the DataverseNL repository, https://doi.org/10.34894/QBBPOK. The repository contains the olink proteomics raw and processed data, associated metadata, and the data analysis scripts used in this study.

## Author contributions

Rianne Rijken: formal analysis, data curation, software, visualization, writing - original draft; Els M Pameijer: investigation, recourses, validation, writing - review & editing; Aafke de Ligt: investigation, resources, writing - review & editing; Marilette Stehouwer: investigation, resources, writing - review & editing; Saskia M Imhof: conceptualization, funding acquisition, supervision, writing - review & editing; Alberta A H J Thiadens: investigation, writing - review & editing; Anneke I den Hollander: investigation, writing - review & editing; Bram Gerritsen: investigation, writing - review & editing; Xuan-Thanh-An Nguyen: investigation, writing - review & editing; Carel B Hoyng: investigation, writing - review & editing; Evianne L de Groot: investigation, writing - review & editing; L. Ingeborgh van den Born: investigation, writing - review & editing; Jeannette Ossewaarde-van Norel: investigation, writing - review & editing; Leonoor I Los: investigation, writing - review & editing; Lude Moekotte: investigation, writing - review & editing; Magda A Smoor: investigation, resources, writing - review & editing; Maria M van Genderen: investigation, writing - review & editing; Ninette H Ten Dam-van Loon: investigation, writing - review & editing; Ramon A C van Huet: investigation, writing - review & editing; Camiel J F Boon: investigation, writing - review & editing; Yvonne de Jong-Hesse: investigation, writing - review & editing; Joke H de Boer: conceptualization, supervision, writing - review & editing; Redmer van Leeuwen: conceptualization, investigation, resources, supervision, writing - review & editing; Jonas J W Kuiper: conceptualization, funding acquisition, methodology, project administration, software, supervision, writing - review & editing. All authors reviewed and approved the article for publication.

## Declaration of Generative AI and AI-assisted technologies

During the preparation of this work the authors used ChatGPT5.5 in the writing process to refine the manuscript and adjust R code. After using this tool, the authors reviewed and edited the content as needed and take full responsibility for the content of the publication.

