## Supplemental Figures for "Cross-Phenotype Plasma Proteomics Reveals Molecular Heterogeneity in Neovascular AMD"

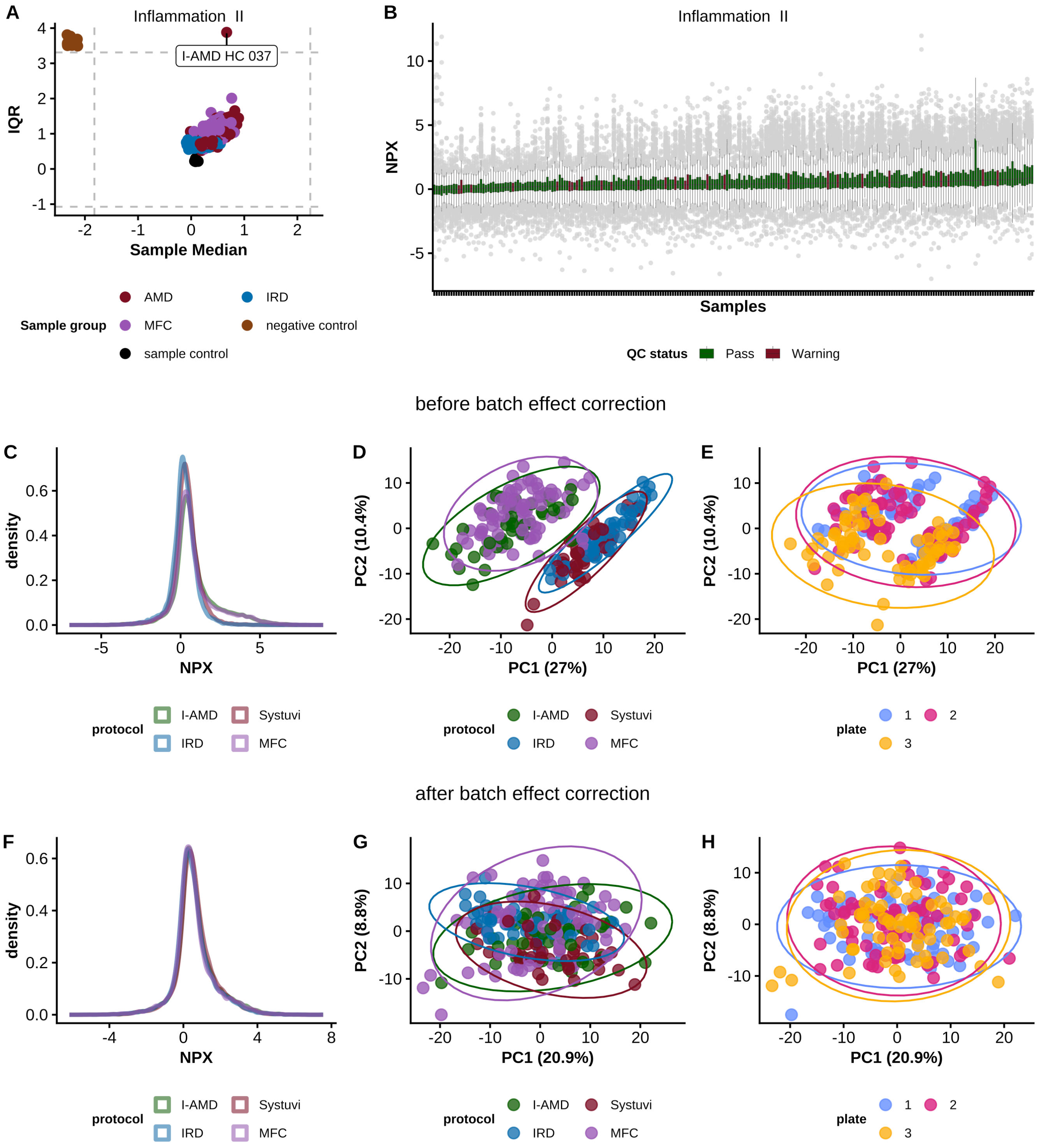


**Supplementary Figure S1: Quality control (QC) of proteomic plasma samples from the Age-Related Macular Degeneration (AMD), Inherited Retinal Diseases (IRD), and Multifocal Choroiditis (MFC) cohorts.** A) The sample median versus the interquartile range (IQR) of Normalized Protein Expression (NPX) values for all plasma samples from the AMD, IRD, and MFC cohorts, along with negative controls and sample controls. The horizontal and vertical grey dashed lines represent an outlier threshold of ±3 standard deviations from the overall sample medians (x-axis) and IQRs (y-axis). Outlier samples from the patient cohorts that exceed this threshold are labeled. B) The distribution of NPX values is shown for all cohort plasma samples in the Inflammation II panel. Boxplots show NPX values (y-axis) for each sample (x-axis). A center line runs through all boxplots, indicating the median. Plasma samples are color-coded based on QC-warning status, where a warning is given if the NPX deviates by more than ±0.3 from the overall sample median on the plate. (C-F) Principal component analysis (PCA) of plasma samples from the patient cohorts, showing the first two principal components. Before batch correction, PCA revealed a freezing-protocol effect (C) and a plate-batch effect (D). After Combat batch correction, PCA demonstrated PCA demonstrated that both the freezing-protocol–related and plate-related batch effects had been effectively removed (E–F). Abbreviations: QC = Quality

Control, AMD = Age-Related Macular Degeneration, IRD = Inherited Retinal Diseases, MFC = Multifocal Choroiditis, IQR = Interquartile Range, NPX = normalized protein expression

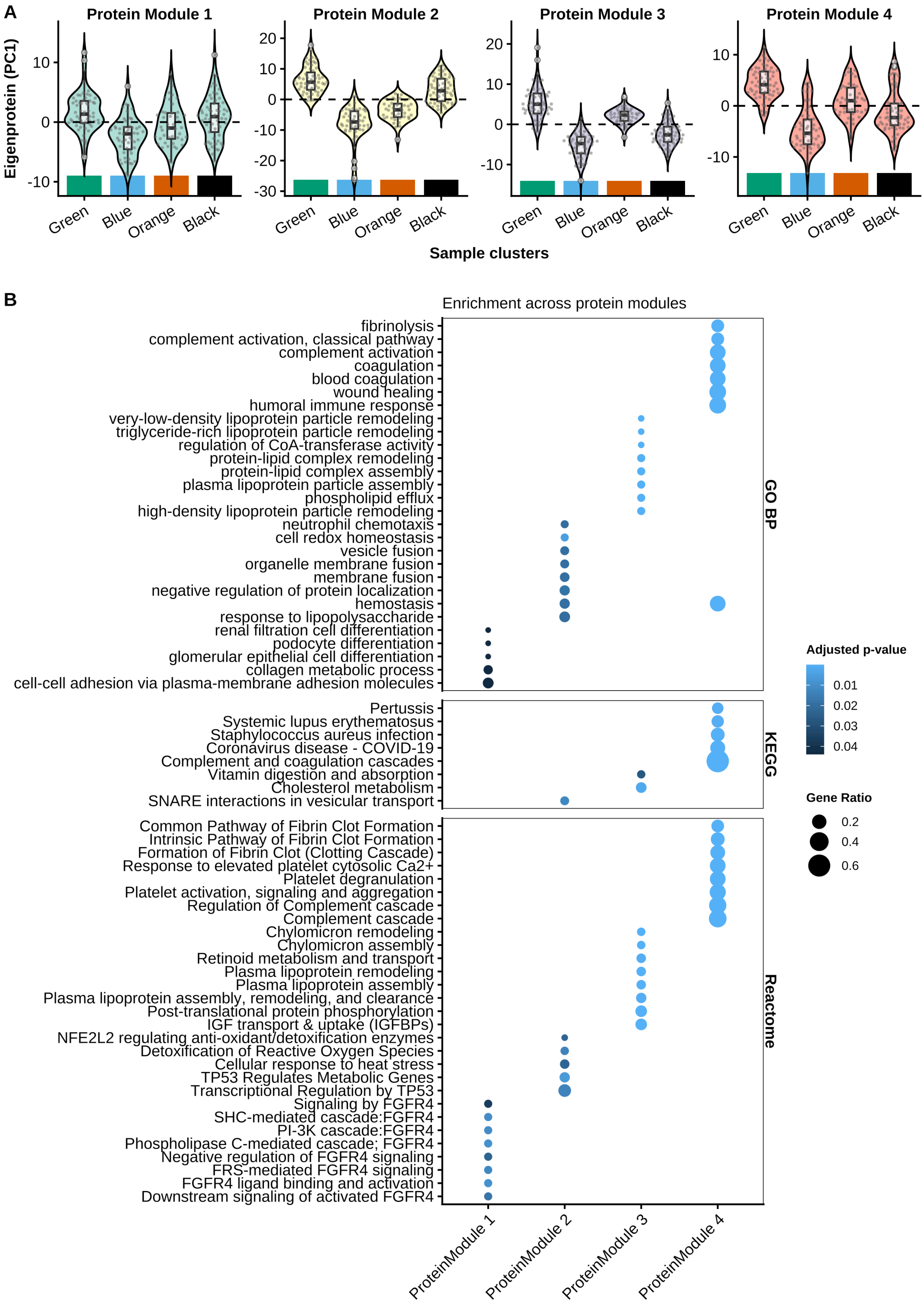
A
Protein Module 1
Protein Module 2
Protein Module 3
Protein Module 4
Eigenprotein (PC1)
Green
Blue
Orange
Black
Sample clusters
B
Enrichment across protein modules
fibrinolysis
complement activation, classical pathway
complement activation
coagulation
blood coagulation
wound healing
humoral immune response
very-low-density lipoprotein particle remodeling
triglyceride-rich lipoprotein particle remodeling
regulation of CoA-transferase activity
protein-lipid complex remodeling
protein-lipid complex assembly
plasma lipoprotein particle assembly
phospholipid efflux
high-density lipoprotein particle remodeling
neutrophil chemotaxis
cell redox homeostasis
vesicle fusion
organelle membrane fusion
membrane fusion
negative regulation of protein localization
hemostasis
response to lipopolysaccharide
renal filtration cell differentiation
podocyte differentiation
glomerular epithelial cell differentiation
collagen metabolic process
cell-cell adhesion via plasma-membrane adhesion molecules
GO BP
Pertussis
Systemic lupus erythematosus
Staphylococcus aureus infection
Coronavirus disease - COVID-19
Complement and coagulation cascades
Vitamin digestion and absorption
Cholesterol metabolism
SNARE interactions in vesicular transport
KEGG
Common Pathway of Fibrin Clot Formation
Intrinsic Pathway of Fibrin Clot Formation
Formation of Fibrin Clot (Clotting Cascade)
Response to elevated platelet cytosolic Ca2+
Platelet degranulation
Platelet activation, signaling and aggregation
Regulation of Complement cascade
Complement cascade
Chylomicron remodeling
Chylomicron assembly
Retinoid metabolism and transport
Plasma lipoprotein remodeling
Plasma lipoprotein assembly
Plasma lipoprotein assembly, remodeling, and clearance
Post-translational protein phosphorylation
IGF transport & uptake (IGFBPs)
NFE2L2 regulating anti-oxidant/detoxification enzymes
Detoxification of Reactive Oxygen Species
Cellular response to heat stress
TP53 Regulates Metabolic Genes
Transcriptional Regulation by TP53
Signaling by FGFR4
SHC-mediated cascade:FGFR4
PI-3K cascade:FGFR4
Phospholipase C-mediated cascade; FGFR4
Negative regulation of FGFR4 signaling
FRS-mediated FGFR4 signaling
FGFR4 ligand binding and activation
Downstream signaling of activated FGFR4
Reactome
ProteinModule 1
ProteinModule 2
ProteinModule 3
ProteinModule 4
Adjusted p-value
0.01
0.02
0.03
0.04
Gene Ratio
0.2
0.4
0.6

**Supplementary Figure S2: Functional characterization of protein modules identified by consensus clustering.** A) Violin plots showing the distribution of module Eigenprotein values (defined as the first principal component of each protein module) across the four sample clusters (*Black*, *Blue*, *Green*, and *Orange*). Each violin represents the variation of the module Eigenprotein among samples within each cluster. Eigenprotein values were centered and scaled, and the horizontal dashed line at zero indicates the reference level of the Eigenprotein scale. Positive Eigenprotein values indicate higher overall expression of proteins in that module within the respective sample cluster. B) Functional pathway enrichment analysis of the four protein modules using Gene Ontology biological processes (GO BP), Kyoto Encyclopedia of Genes and Genomes (KEGG) pathways, and Reactome pathways. The top significantly enriched pathways per protein module and pathway database are shown, with circle size corresponding to the proportion of module proteins annotated to each pathway and color representing the adjusted p-value.Abbreviations: PC1 = First principal component; GO BP = Gene Ontology Biological Process; KEGG = Kyoto Encyclopedia of Genes and Genomes

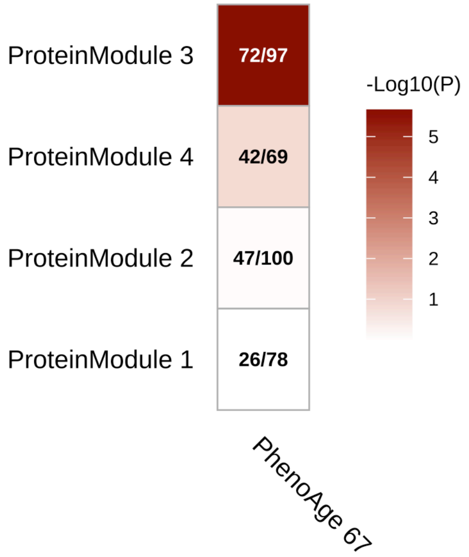


**Supplementary Figure S3: Enrichment analysis of biological age signature (PhenoAge 67) established in >50,000 individuals of the UK Biobank** (from Ma et al. 2026)[25]. Enrichment for PhenoAge 67 signature was assessed using a one-sided hypergeometric over-representation test. using the full set of module proteins as the background. *P*-values were adjusted for multiple testing using the Benjamini–Hochberg procedure. Cell annotations indicate the number of overlapping proteins relative to total module size. Abbreviations: PhenoAge = phenotypic age; *P* = p-value
